# TOWARD A COVID-19 SCORE-RISK ASSESSMENTS AND REGISTRY

**DOI:** 10.1101/2020.04.15.20066860

**Authors:** M Cristina Vazquez Guillamet, Rodrigo Vazquez Guillamet, Andrew A. Kramer, Paula M. Maurer, Gregory A. Menke, Cherie L. Hill, William A. Knaus

## Abstract

**Importance:** Critical care resources like ventilators, used to manage the current COVID-19 pandemic, are potentially inadequate. Established triage standards and guidelines may not contain the most appropriate severity assessment and outcome prediction models.

**Objectives:** Develop a draft pandemic specific triage assessment score for the current COVID-19 pandemic. Design a website where initial Toward a COVID-19 Scores (TACS) can be quickly calculated and used to compare various treatment strategies. Create a TACS Registry where data and outcomes for suspected and confirmed COVID-19 patients can be recorded. Use the TACS Registry to develop an influenza epidemic specific database and score for use in future respiratory based epidemics.

**Design, Setting, Participants:** Retrospective analysis of 3,301 ICU admissions with respiratory failure admitted to 41 U.S. Intensive Care Units from 2015-19. Independent external validation on 1,175 similar ICU Admissions using identical entry criteria from Barnes Jewish Hospital (BJH), Washington University from 2016-2019.

**Main Outcomes:** TACS was created with 16 readily available predictive variables for risk assessment of hospital mortality 24 hours after ICU Admission and the need for prolonged assisted mechanical ventilation (PAMV) (> 96 hours) at 24- and 48-hours post ICU admission.

**Results:** TACS achieved an Area Under the Curve (AUC) for hospital mortality after 24 hours of 0.80 in the development dataset; 0.81 in the internal validation dataset. At a probability of 50% hospital mortality, positive predictive value (PPV) was 0.55, negative predictive value (NPV) 0.89; sensitivity 22%, specificity 97%.

For PAMV after 24 hours, the AUC was 0.84 in the development dataset, 0.81 in the validation dataset. For PAMV after 48 hours, the AUC was 0.82 in the development dataset, 0.78 in the validation dataset.

In the external validation the AUC for TACS was 0.76 +/- 0.024. We launched a website that is scaled for mobile device use (https://covid19score.azurewebsites.net/) that provides open access to a user-friendly TACS Calculator for all predictions. We also designed a voluntary TACS Registry for collection of data and outcomes on ICU Admissions with COVID-19.

**Conclusions and Relevance:** Toward a COVID-19 score is a starting point for an epidemic specific triage assessment that could be used to evaluate various approaches to treatment. The TACS Registry provides the ability to establish a respiratory specific outcomes database that can be used to create a triage approach for future such pandemics.

**Key Points:** *Question:* Can a rapid epidemic specific risk assessment severity score and data and outcome repository be constructed in the midst of a pandemic.

*Findings:* Using development and validation datasets with ICU admissions similar to those developing COVID-19, developed an initial Toward a COVID-19 Score that could be used to compare various treatment approaches. Also launched an online facilitated data collection and outcome assessment registry for collection of a pandemic specific database so a new triage score could be created for use in the next pandemic.

*Meaning:* In the midst of a pandemic rapid development of an epidemic specific triage score and a data registry for the creation of a new score for use in future pandemics appears feasible.

## Introduction

Critical care resources like ventilators, used to manage the COVID-19 pandemic, are reported inadequate worldwide ^1^. In the US established triage standards, such as Ventilator Allocation Guidelines developed by New York State ^2^, by Johns Hopkins ^3^, and Pennsylvania^4^, have been recently endorsed^5^. These represent substantial efforts to design an ethical allocation system for a range of disaster scenarios.

One component of these may not be optimal in the current pandemic: sole reliance on the Sequential Organ Failure Assessment (SOFA) score for severity measurement^6^. SOFA was developed primarily for use in hospitalized sepsis. It has one respiratory measurement: the PaO2/FIO2 ratio. The other five measures document deterioration in other organ systems. Published data consistently report many COVID-19 patients did not develop multi organ failure during the Intensive Care Unit stay^7–9^. US and European colleagues treating COVID-19 patients confirm that many deaths are from primary respiratory, not multiple system, failure ^10^. SOFA scores for triage in a COVID-19 pandemic may then not discriminate who would benefit from intubation and only be useful late in a patient’s course, after they have received multiple days or weeks of mechanical ventilation. More so, since COVID-19 patients will presumably have lower SOFA score at the time of intubation compared to septic patients they will be allocated a ventilator even though current articles report a significantly higher mortality^7,11^.

Our objective was to begin a process whereby a primarily respiratory based severity assessment measure that also incorporated other known patient characteristics associated with this COVID-19 epidemic and patient outcomes could be established. More importantly, we sought to establish a COVID-19 Registry so a database that could be used to develop a new score for future pandemics.

### Summary of Methods

We designed and validated an initial TACS based on 3,301 adult patients with a clinical diagnosis of respiratory distress admitted to 41 ICUs in the US from 2015-19.

We performed an additional external validation of the TACS in 1,175 ICU Admissions to Barnes Jewish Hospital (BJH), Washington University from 2016-2019.

### Statistical Methods

The statistical models were initially developed and internally validated on admissions from 1/1/2015 through 9/30/2015 in Medical Decision Network’s, Charlottesville, Va. (MDN) Phoenix ICU database. Phoenix provides data critical to research efforts such as the development of TACS as well as providing insight and understanding to clinicians and administrators making front line decisions that impact patient care.

The dataset provided by MDN for the development of TACS consists of 3,301 de-identified admissions admitted to 41 adult ICUs. To qualify, an ICU had to send electronically captured vital signs to MDN. These ICUs were situated in the Southeast, Northeast, Mid-Atlantic and Pacific West regions. Patients had to be 18 years and above and have one of the following admitting diagnoses: asthma, COPD, pneumonia (bacterial, viral, or parasitic), pulmonary edema, respiratory arrest, restrictive lung disease (fibrosis, sarcoidosis), sleep apnea, or hemorrhage/hemoptysis.

Variables considered as predictors were organized as demographics, comorbidities and, pulmonary physiology parameters. Vital signs and labs were formatted dependent on methodology taken from the Acute Physiology, Age, and Chronic Health Evaluation (APACHE) system ^12–14^. The vital signs data were aggregated into minimum and maximum value within the first 24 hours after admission (e.g. max_hr24) and within 24 and 48 hours (e.g. max_temp48). The same was carried out for albumin and ph. Based on clinical knowledge the following vitals and labs were entered into the multivariable models:

Maximum heart rate

Maximum respiratory rate

Minimum mean arterial pressure

Maximum temperature

Minimum albumin

Minimum pH

Missing values for albumin were imputed as the median value of 2.80, while missing pH values were imputed with the median value of 7.39. None of the min/max vital signs had missing values.

Along with vital signs and the two lab values, a patient’s age, gender = female, square root of the length of time between hospital admission and ICU admission, (This was done to reduce the effect of extreme (long stay) patients),. minimum PaO2:FiO2 ratio during the first 24 hours after admission, maximum Glasgow Coma Score during the first 24 hours after admission, the presence or absence of the following comorbidities: COPD, congestive heart failure, chronic alcoholism, body mass index > 30, and chronic tobacco use. When arterial blood gases where not available, PaO2/FiO2 ratio was imputed by the APACHE methodology, = 80/.0.21. Another variable combined seven comorbidities indicative of immunosuppression was derived by a patient having one or more of the following comorbidities based on the APACHE methodology: AIDS, cirrhosis, leukemia, lymphoma, metastatic cancer, hepatic failure, and diabetes. Finally, interaction terms were created for age with COPD, congestive heart failure, body mass index, and male gender. All of these variables were chosen based on clinical expertise, the emerging literature describing COVID-19 cases, and descriptions provided by front-line practitioners and no addition or removal of variables was performed based on statistical significance. To account for the effects of when a patient was placed on mechanical ventilation, a variable called “initMV” was computed as the duration between ICU admission and hospital admission. If greater than 1.00 days, this value was set equal to zero for the 24-hour model (see below); for the 48-hour models (see below) initMV was truncated at 2.00 days.

The MDN data set was divided randomly 2:1 into development and validation data sets. A logistic regression procedure was carried on the development data set. Variables remained in the model regardless of statistical significance, as they were deemed clinically important. Model accuracy was determined by the area under the receiver operating characteristic curve ^15^and the ratio of observed to predicted outcomes. The coefficients from the development data set model were then fed into the validation data set, and the same statistics on this group of patients were obtained. Finally, a cut-point probability of 0.50 was used to calculate the positive predictive value, negative predictive value, sensitivity and specificity of the model. There were three outcomes for which the above procedures were carried out: hospital mortality given labs and vital signs at 24 hours, PAMV given labs and vital signs at 24 hours, and PAMV given labs and vital signs at 48 hours. For the outcome mortality at 24 hours, only a patient’s first ICU admission was included (to avoid counting one patient twice). When developing the models with PAMV as the outcome, all ICU admissions were included, except patients not in the ICU after 24 and 48 hours respectively were excluded.

For external validation, the 24h TACS mortality model was replicated in the Barnes Jewish Hospital dataset. The patients were selected with the same entry criteria as our developmental and validation datasets (see statistical appendix).

## Results

Table 1 provides demographic and clinical characteristics of the MDN and Washington University -BJH datasets, respectively. All values given are the mean or percentage, depending on the variable’s distribution. The MDN data set had patients who were older, had less immunocompromised comorbidities, longer previous length of stay, ICU length of stay, and a higher percentage of patients with prolonged acute mechanical ventilation (“PAMV”; duration on ventilation > 96 hours) (14).

**Table 1.** Demographic and clinical characteristics of the patients in the MDN and WU datasets. Continuous variables have mean and the Intra-quartile range.

|  | MDN Dataset<br>n=3,177 | WU Dataset n = 1,175 |
| --- | --- | --- |
| Age (yrs.) | 66.1 [57, 77] | 59.3 [52, 69] |
| Gender = Female | 52.9% | 53.2% |
| Patients with Immunocompromised Comorbidities | 21.0% | 30.0% |
| Hospital Mortality | 13.4% | 13.1% |
| Mechanically Ventilated on Day 1 | 35.4% | 34.2% |
| Square root of Previous Length of Stay in the Hospital (days) | 0.77 [0.41, 0.93] | 0.33 [0.26, 0.44] |
| ICU Length of Stay (days) | 3.97 [1.27, 4.88] | 2.92 [1.24, 3.63] |
| Duration of Mechanical Ventilation (days) | 2.70 [0.75, 3.46] | 2.31 [0.62, 3.00] |
| Prolonged Acute Mechanical Ventilation | 7.3% | 4.5% |

MDN = Medical Decision Network
WU = Washington University

Tables 2a, 2b, and 2c give each model’s coefficients, standard error, and p-value. Figures 1a, 1b, and 1c show each model’s calibration curve: observed vs. predicted outcome over deciles of predicted outcome within the MDN dataset.

**Table 2a.**
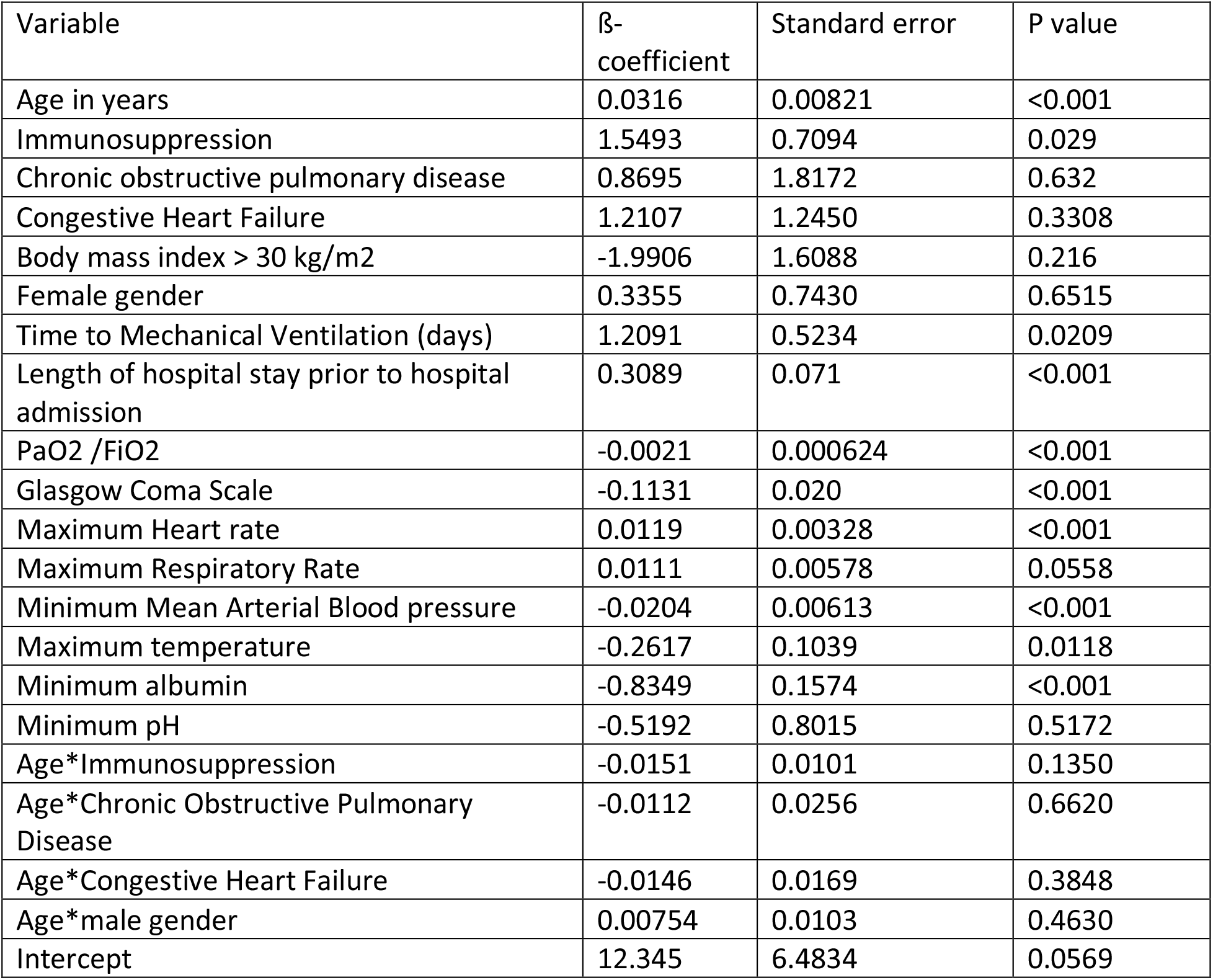
In-hospital mortality based on variables collected during the first 24-hours within the MDN dataset

| Variable | β-coefficient | Standard error | P value |
| --- | --- | --- | --- |
| Age in years | 0.0316 | 0.00821 | <0.001 |
| Immunosuppression | 1.5493 | 0.7094 | 0.029 |
| Chronic obstructive pulmonary disease | 0.8695 | 1.8172 | 0.632 |
| Congestive Heart Failure | 1.2107 | 1.2450 | 0.3308 |
| Body mass index > 30 kg/m<sup>2</sup> | -1.9906 | 1.6088 | 0.216 |
| Female gender | 0.3355 | 0.7430 | 0.6515 |
| Time to Mechanical Ventilation (days) | 1.2091 | 0.5234 | 0.0209 |
| Length of hospital stay prior to hospital admission | 0.3089 | 0.071 | <0.001 |
| PaO<sub>2</sub> /FiO<sub>2</sub> | -0.0021 | 0.000624 | <0.001 |
| Glasgow Coma Scale | -0.1131 | 0.020 | <0.001 |
| Maximum Heart rate | 0.0119 | 0.00328 | <0.001 |
| Maximum Respiratory Rate | 0.0111 | 0.00578 | 0.0558 |
| Minimum Mean Arterial Blood pressure | -0.0204 | 0.00613 | <0.001 |
| Maximum temperature | -0.2617 | 0.1039 | 0.0118 |
| Minimum albumin | -0.8349 | 0.1574 | <0.001 |
| Minimum pH | -0.5192 | 0.8015 | 0.5172 |
| Age*Immunosuppression | -0.0151 | 0.0101 | 0.1350 |
| Age*Chronic Obstructive Pulmonary Disease | -0.0112 | 0.0256 | 0.6620 |
| Age*Congestive Heart Failure | -0.0146 | 0.0169 | 0.3848 |
| Age*male gender | 0.00754 | 0.0103 | 0.4630 |
| Intercept | 12.345 | 6.4834 | 0.0569 |

**Table 2b.** Duration of ventilation greater than 96h based on variables collected during the first 24 hours within the MDN dataset.

| Variable | β-coefficient | Standard error | P value |
| --- | --- | --- | --- |
| Age in years | -0.00603 | 0.00689 | 0.3817 |
| Immunosuppression | -0.0207 | 0.9044 | 0.9817 |
| Chronic obstructive pulmonary disease | -2.2227 | 2.7307 | 0.4157 |
| Congestive Heart Failure | 1.4462 | 1.3105 | 0.2698 |
| Body mass index > 30 kg/m<sup>2</sup> | 0.9181 | 1.3878 | 0.5082 |
| Female gender | 0.4730 | 0.1868 | 0.0113 |
| Time to Mechanical Ventilation (days) | 1.9293 | 0.5104 | 0.0002 |
| Length of hospital stay prior to hospital admission | 0.0909 | 0.0797 | 0.2537 |
| PaO<sub>2</sub> /FiO<sub>2</sub> | -0.00534 | 0.000808 | <0.001 |
| Glasgow Coma Scale | -0.1840 | 0.0258 | <0.001 |
| Maximum Heart rate | 0.00156 | 0.00437 | 0.7213 |
| Maximum Respiratory Rate | 0.00157 | 0.00761 | 0.7213 |
| Minimum Mean Arterial Blood pressure | -0.00828 | 0.00793 | 0.2960 |
| Maximum temperature | 0.1679 | 0.1249 | 0.1788 |
| Minimum albumin | -0.5262 | 0.1955 | 0.0071 |
| Minimum pH | 1.5345 | 0.9391 | 0.1023 |
| Age*Immunosuppression | -0.00081 | 0.0134 | 0.9516 |
| Age*Chronic Obstructive Pulmonary Disease | 0.0244 | 0.0384 | 0.5258 |
| Age*Congestive Heart Failure | -0.0163 | 0.0188 | 0.3853 |
| Age*Body mass index>30kg/m<sup>2</sup> | -0.0131 | 0.0213 | 0.5394 |
| Intercept | -15.1693 | 7.6738 | 0.0481 |

**Table 2c.** Duration of mechanical ventilation greater than 96-hours using data collected from 24 to 48 hours within the MDN dataset.

| Variable | β-coefficient | Standard error | P value |
| --- | --- | --- | --- |
| Age in years | -0.00485 | 0.00754 | 0.5202 |
| Immunosuppression | 0.1970 | 1.0454 | 0.8505 |
| Chronic obstructive pulmonary disease | -1.3560 | 2.8911 | 0.639 |
| Congestive Heart Failure | 2.7507 | 1.5968 | 0.085 |
| Body mass index > 30 kg/m<sup>2</sup> | 0.6144 | 0.4964 | 0.6814 |
| Female gender | 0.4552 | 0.2054 | 0.0267 |
| Time to Mechanical Ventilation (days) | 1.8672 | 0.2905 | <0.0001 |
| Length of hospital stay prior to hospital admission | -0.0526 | 0.1253 | 0.6744 |
| PaO<sub>2</sub> /FiO<sub>2</sub> | -0.00441 | 0.000857 | <0.0001 |
| Glasgow Coma Scale | -0.1918 | 0.029 | <0.0001 |
| Maximum Heart rate | -0.00207 | 0.00489 | 0.6715 |
| Maximum Respiratory Rate | -0.00342 | 0.00741 | 0.6443 |
| Minimum Mean Arterial Blood pressure | 0.00329 | 0.0082 | 0.688 |
| Maximum temperature | -0.0366 | 0.1445 | 0.8002 |
| Minimum albumin | 0.2362 | 0.2482 | 0.3414 |
| Minimum pH | -0.5081 | 0.9529 | 0.5939 |
| Age*Immunosuppression | -0.00579 | 0.0156 | 0.7099 |
| Age*Chronic Obstructive Pulmonary Disease | 0.0129 | 0.0410 | 0.7533 |
| Age*Congestive Heart Failure | -0.0347 | 0.0227 | 0.1258 |
| Age*Body mass index>30kg/m<sup>2</sup> | -0.00467 | 0.0229 | 0.8382 |
| Intercept | 0.7683 | 0.5908 | 0.1934 |

**Figure 1a.**
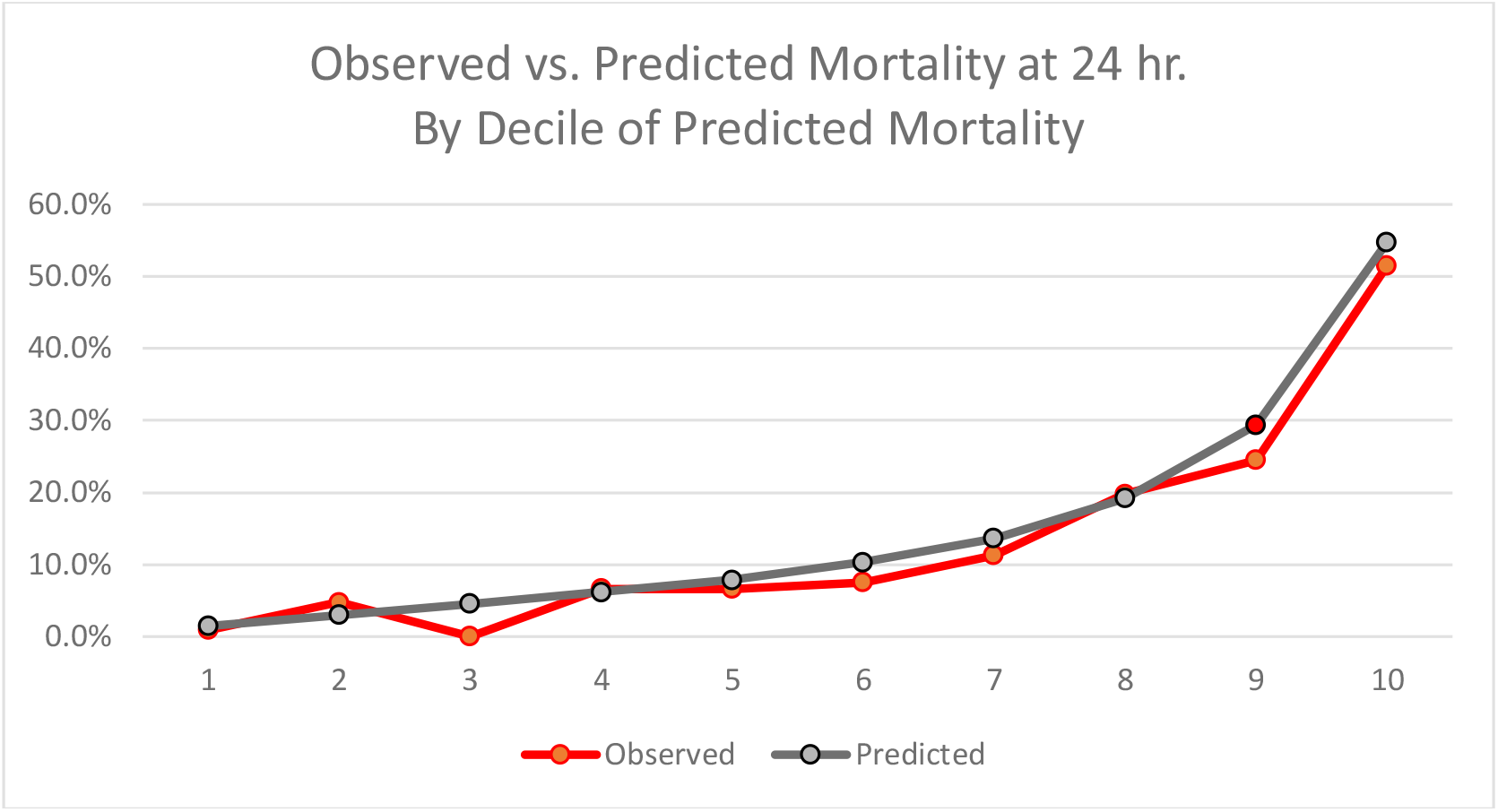
24 Hour Model: Observed vs. Predicted Hospital Mortality Across Deciles of Predicted Mortality within the MDN Dataset (N= 3,301) for TACS.

**Figure 1b.**
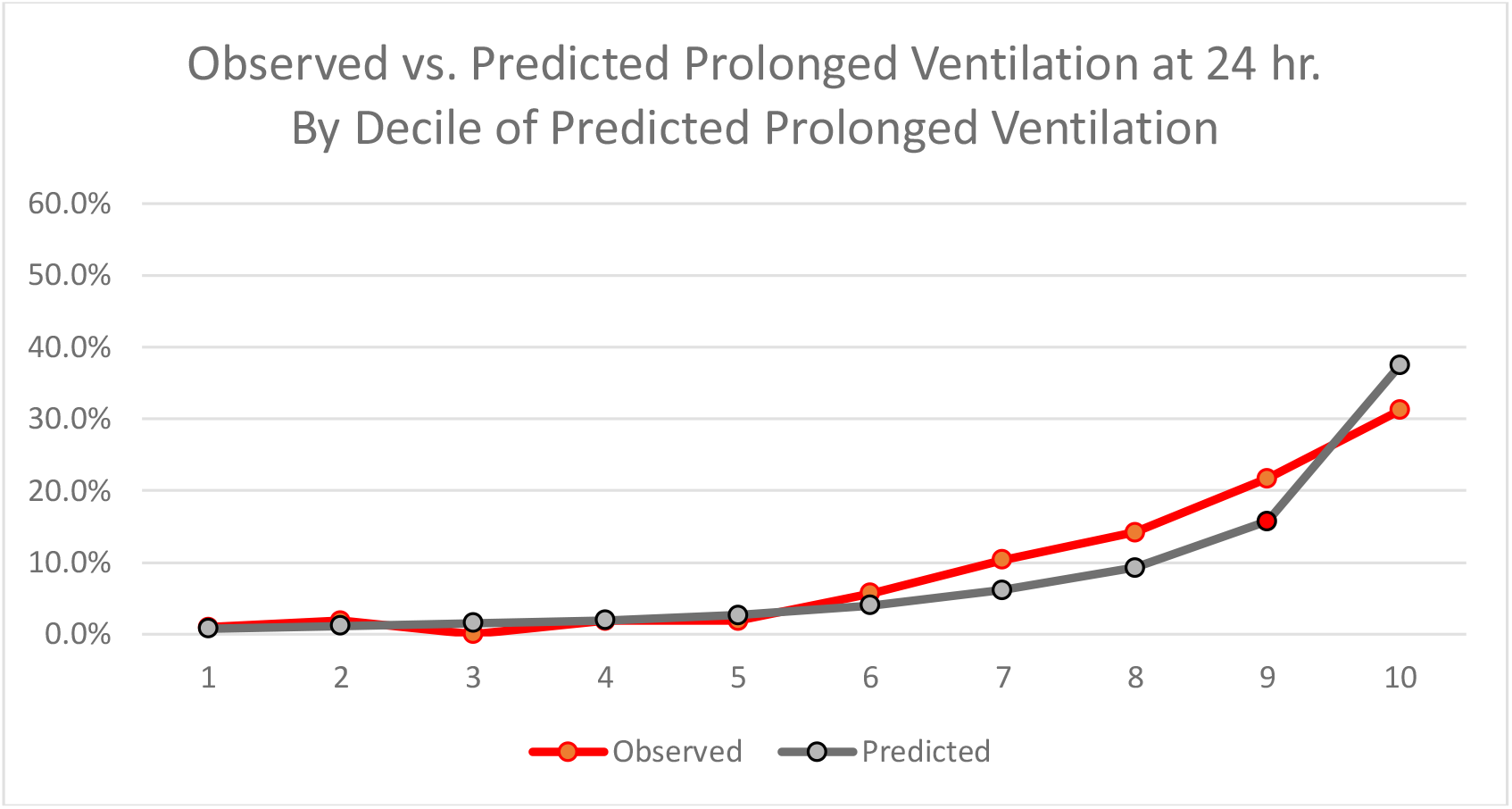
24 Hour Model: Observed vs. Predicted Prolonged Ventilation across Deciles of Predicted Prolonged Ventilation within the MDN Dataset (N=3,301) for TACS.

**Figure 1c.**
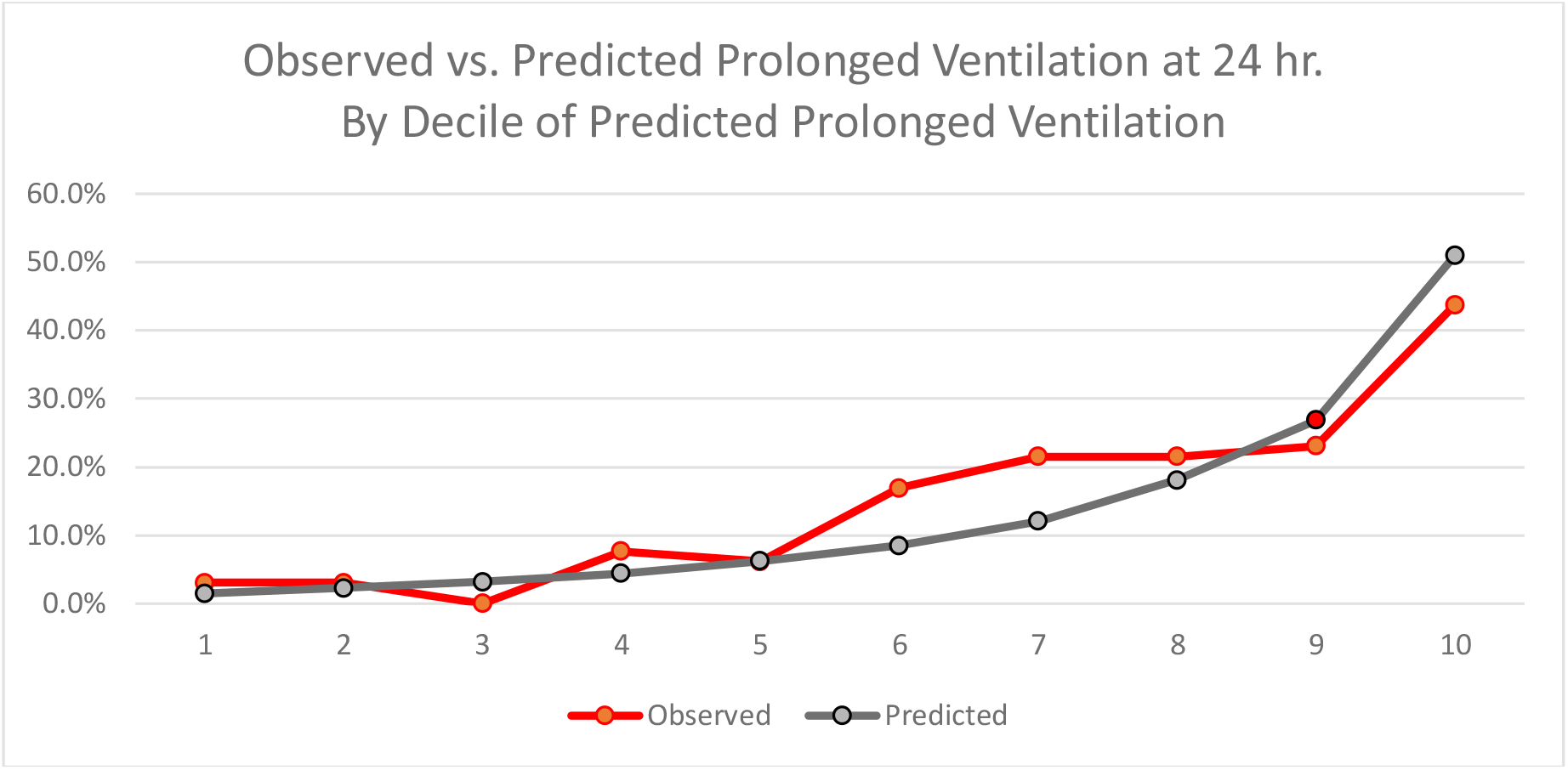
48 Hour Model: Observed vs. Predicted Prolonged Ventilation across Deciles of Predicted Prolonged Ventilation within the MDN Dataset (N-3,301) for TACS.

Table 3 displays the performance for predicting hospital mortality within the first 24 hours of ICU admission and prolonged ventilation within the first 24 and 48 hours. TACS achieved an Area Under the Curve (AUC) for predicting hospital mortality after 24 hours of ICU treatment of 0.80 in the development dataset; 0.81 in the internal validation dataset. At a probability of 50% for hospital mortality, the positive predictive value (PPV) is 0.55, negative predictive value (NPV) 0.89; sensitivity 22%, specificity 97%.

**Table 3.** Results from the multivariable TACS models to predict hospital mortality at 24 hours after admission, and prolonged acute mechanical ventilation at 24 and 48 hours, respectively.

**Hospital Mortality Prediction at 24 hours post-admission**
| Data Set | N | observed % | predicted % | SMR | p-value | AU-ROC | PPV | NPV |
| --- | --- | --- | --- | --- | --- | --- | --- | --- |
| Development | 2122 | 13.5% | 13.5% | 1.00 | >0.10 | **0.80** | n/a | n/a |
| Validation | 1055 | 13.2% | 14.8% | 0.89 | 0.09 | **0.81** | 0.55 | 0.89 |

**Prolonged Acute Mechanical Ventilation ( >96 hours) Prediction at 24 hours post admission**
| Data Set | N | observed % | predicted % | SMR | p-value | AU-ROC | PPV | NPV |
| --- | --- | --- | --- | --- | --- | --- | --- | --- |
| Development | 2167 | 7.3% | 7.3% | 1.00 | >0.10 | **0.84** | n/a | n/a |
| Validation | 1063 | 9.0% | 8.2% | 1.11 | >0.10 | **0.81** | 0.38 | 0.92 |

**Prolonged Acute Mechanical Ventilation ( >96 hours) Prediction at 48 hours post admission**
| Data Set | N | observed % | predicted % | SMR | p-value | AU-ROC | PPV | NPV |
| --- | --- | --- | --- | --- | --- | --- | --- | --- |
| Development | 1169 | 12.1% | 11.7% | 1.04 | >0.10 | **0.82** | n/a | n/a |
| Validation | 619 | 14.5% | 12.6% | 1.15 | > 0.10 | **0.78** | 0.583 | 0.87 |

SMR = Standardized Mortality Ratio (observed % / predicted %). An SMR > 1.0 indicates more outcomes ( mortalities or prolonged ventilations ) than expected based on patient characteristics. An SMR < 1.0 indicates fewer observed outcomes than estimated.
p-value is the probability under the Chi-square distribution that the test statistic $\Sigma(\text{observed} (0,1) - \text{predicted})^2 / \Sigma(\text{observed} (0, 1) * \text{predicted} )$ exceeds 3.842.
AU-ROC = area under the Receiver Operating Characteristics curve. AUC ranges in value from 0 to 1. A model whose predictions are 100% wrong has an AUC of 0.0; one whose predictions are 100% correct has an AUC of 1.0. A value of 0.5 means a prediction equal to a coin flip or chance.
PPV = Number of True Positives/Number of Positive Predictions
Positive Predictive Value is probability that subjects with a positive prediction truly have the outcome ( hospital mortality and/or prolonged ventilation).
NPV = Number of True Negatives/ Number of Negative Predictions
Negative Predictive Value is the probability that subjects with a negative prediction truly don't have the outcome (hospital mortality and/or prolonged ventilation).

For prediction of the need for prolonged mechanical ventilation after 24 hours of ICU treatment, the TACS AUC was 0.84 in the development dataset, 0.81 in the validation dataset. For predictions of prolonged mechanical ventilation after 48 hours of ICU treatment, the TACS AUC was 0.82 in the development dataset, 0.78 in the validation dataset.

We also performed an external validation of the TACS 24-hour mortality model on 1,1175 ICU patients to Washington University/BJH treated between 2016-19. Mortality prediction at 24 hours the TACS AUC was 0.76 +/- 0.024.

## DISCUSSION

We have developed an initial model of a respiratory oriented Toward a COVID 19 Score designed to be useful in possible triage decisions and to compare outcomes from various treatment approaches in the current pandemic. It was designed specifically with this pandemic in mind. We used the early reports from China and discussions with front line practitioners treating COVID-19 patients as the basis for selecting predictor variables. While it focuses on respiratory physiology parameters, it also has seven possible choices for immunosuppression among other pre-existing conditions, as these are well-documented and very important risk factors. The final model is broad enough to include all possible current and future COVID trajectories but also narrow enough to focus on death from respiratory causes such as Adult Respiratory Distress Syndrome (ARDS).

While previous attempts at developing an ARDS mortality prediction models were inferior to more general multi organ failure scores such as APACHE, those models combined a very heterogenous ARDS population ^16^The model relied on age, bilirubin, hematocrit and net volume balance over 24hrs. Pneumonia severity prediction tools including respiratory parameters have maintained their predictive performance (curb-65, psi). Since current reports describe COVID-19 pneumonia as predominantly a single organ failure we decide to focus on respiratory parameters.

A precursory study by Luo et al described that direct ARDS had fewer organ failures and different mortality predictors than indirect ARDS (e.g. non-pneumonia sepsis) despite similar mortality rates. The lung injury scores correlated with mortality in direct ARDS but not in indirect ARDS ^17^. In our study we also focused on lung physiology parameters along with variables that have been described in COVID reports.

In rationing situations, the main use for a risk stratification score would be to discriminate survivors and non-survivors among the critically ill so patients more likely to survive will be allocated a ventilator. A secondary use would be serial measurements to assess the trajectory of intubated patients and predict patients who will require prolonged mechanical ventilation. A third use would be to compare outcomes from various treatment strategies. We initially focused on these objectives. While earlier reports from China showed that higher SOFA scores were associated with higher mortality, all hospitalized patients were included ^18^. More than half of the patients were admitted to the general wards therefore comparing non-critically ill and critically ill patients. This comparison will not be relevant in crisis situations when the selection criteria will be applied solely to critically ill patients. This is why in our model, we included patients who had an ICU stay. Other studies have used SOFA in critically ill patients but with very small samples sizes, only 9 outcomes thus questioning the validity of the analysis ^19^. When analyzing 52 critically ill patients, Yang et al found the initial 24hr SOFA score was 4 in survivors compared to 6 in non-survivors ^19^. The traditional cutoff for SOFA is < 6 which correlates with a mortality of 10% ^20^. The findings by Yang et al reinforce the concept that early SOFA will not discriminate between survivors and non-survivors as most patients present to the ICU in single organ failure. In the study by Yang et al, the in-hospital mortality was 61.5% although the SOFA score in non-survivors predicted a mortality below 10%.

The final TACS 24 hour mortality model displayed good discrimination within the independent external Washington University dataset. We anticipate, however, that when the model is applied to a COVID-19 ICU cohort, the observed hospital death rates will be much higher than the 13% −14 % found in our derivation, validation, and external datasets ^9^. Under these circumstances, the model’s PPV at higher probabilities will be strengthened. In a pandemic, patients with the highest likelihood of survival should have priority access to therapy. It is our expectation that TACS should be able to identify early the patients more likely to benefit from life support when resources are limited.

## LIMITATION

The major limitation of TACS is that it has not been applied to patients with a confirmed COVID 19 diagnosis. This is a major question. But, waiting for a substantial database of COVID-19 cases with confirmed outcomes in this country, meant there will have been many deaths and many decisions made with only a potentially adequate severity/outcome estimate, SOFA, as part of the decision making. We have therefore chosen to release it now with the intent of being able to acquire new COVID 19 cases as quickly as possible and improving upon this base. We will commit to providing new data on TACS’s performance with COVID-19 patients as soon as possible. We also acknowledge that we will need to create outcome predictions for later in the ICU stay. These will be a major focus of the TACS Registry along with development of a database in order to create a new revised score for use in the next such pandemic.

## WEBSITE AND VOLUNTARY EPIDEMIC SPECIFIC REGISTRY

We have launched a website with mobile access (https://covid19score.azurewebsites.net/) that give open access to a user-friendly TACS Calculator for all predictions. Because TACS was developed on non-COVID cases, we believe it can be applied to other causes of respiratory failure as appropriate. A voluntary registry option reporting subsequent patient outcomes will enable us to further validate and make improvements in the score and our understanding of COVID-19. The TACS registry requests registration and the recording of patient outcomes (hospital mortality, hospital LOS, and duration of mechanical ventilation). It also enables data to be recorded over the course of the hospital stay. Collection of a pandemic specific database will allow use to create a new respiratory based resource for use in future such pandemics.

### Conclusion

The Toward A COVID-19 score is designed to be used within an overall rationing approach when critical care resources are scarce (2-5). It provides an earlier and conceptually superior prediction of outcomes compared to SOFA. It also can be used to compare outcomes from various approaches to treatment of COVID-19 ICU admissions. Finally, we have established a COVID-19 Registry to collect and store data and outcomes on patients treated during this pandemic. TACS Registry data will hopefully be available to develop a new score for use in the next such pandemic (21).

## Data Availability

The full equations are published in the manuscript. The accompanying website and registry are open to all users.

https://covid19score.azurewebsites.net/

